# Global estimates of lives and life-years saved by COVID-19 vaccination during 2020-2024

**DOI:** 10.1101/2024.11.03.24316673

**Authors:** John P.A. Ioannidis, Angelo Maria Pezzullo, Antonio Cristiano, Stefania Boccia

## Abstract

**Importance:** Estimating global lives and life-years saved is important to put into perspective the benefits of COVID-19 vaccination. Prior studies have focused mainly on the pre-Omicron period or only on specific regions, lack crucial life-year calculations, and often depend on strong modeling assumptions with unaccounted uncertainty.

**Observations:** We aimed to calculate the lives and life-years saved by COVID-19 vaccination worldwide from the onset of the vaccination campaigns and until October 2024. We considered different strata according to age; community-dwelling and long-term care residence status; pre-Omicron and Omicron periods; and vaccination before and after a SARS-CoV-2 infection. In the main analysis, 2.533 million deaths were averted (1 death averted per 5,400 vaccine doses administered). Eighty-two percent were among people vaccinated before any infection, 57% were in the Omicron period, and 90% pertained to people 60 years and above. Sensitivity analyses suggested 1.4 to 4.0 million lives saved. Some sensitivity analyses showed preponderance of the benefit during the pre-Omicron period. We estimated 14.8 million life-years saved (1 life-year saved per 900 vaccine doses administered). Sensitivity range was 7.4-23.6 million life-years. Most life-years saved (76%) were in people over 60 years old, but long-term care residents contributed only 2% of the total. Children and adolescents (0.01% of lives saved and 0.1% of life-years saved) and young adults 20-29 years old (0.07% of lives saved and 0.3% of life-years saved) had very small contributions to the total benefit.

**Conclusions and relevance:** Based on a number of assumptions, these estimates are substantially more conservative than previous calculations focusing mostly on the first year of vaccination, but they still clearly demonstrate a major overall benefit from COVID-19 vaccination during 2020-2024. The vast majority of benefit in lives and life-years saved was secured for a portion of the elderly minority of the global population.

## INTRODUCTION

The development and wide implementation of COVID-19 vaccines are widely considered major successes for biomedical research and public health.^1,2^ It is important to estimate the number of lives saved by COVID-19 vaccination worldwide since their introduction. Previous efforts to estimate deaths averted by COVID-19 vaccines used epidemic modeling or counterfactuals from surveillance data.^3–6^ Models may give unreliable results, depending on assumptions.^7,8^ Moreover, most previous publications addressed the pre-Omicron period.^3,4^ The few that have included Omicron period data^5,6^ focused on specific regions and have not calculated probable life-years saved. Life-year estimates are pivotal in decision-making.

Here we estimate both lives and life-years saved by COVID-19 vaccination worldwide until October 2024, separating different age strata, community-dwelling and long-term care populations, pre-Omicron and Omicron periods, and populations vaccinated before or after SARS-CoV-2 infection.

## METHODS

### Outline of calculations

We consider strata based on age and long-term care residence status. Furthermore, we separate the period until November 2021 (pre-Omicron) and the subsequent period (Omicron); and people vaccinated before any SARS-CoV-2 infection from those vaccinated after previous infection. Stratifications are important because the infection fatality rate (IFR) varies markedly across strata.

The number of lives saved in each stratum i is the product of the number who would have died absent vaccination, and vaccine effectiveness (VE_i_) for mortality. The number of people who would have died is the product of the total stratum population N_i_, the proportion who would have been infected PI*_i_ (absent vaccination), and the respective IFR_i_:

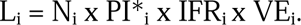

Total lives saved are

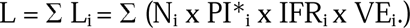

Similarly, life-years saved LY_i_ are proportional to L_i_, the stratum-specific life expectancy (LE) LE_i_, and to a factor f_i_ that denotes how LE of those who died may have differed versus general population LE; f takes smaller values when those who die are in worse health than the respective same-stratum general population. Thus:

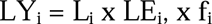

Total life-years saved are

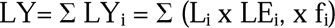

We first calculate the benefits for people vaccinated before any SARS-CoV-2 infection. For those first vaccinated after having at least one SARS-CoV-2 infection, we then assume that PI*_i_ x IFR_i_ x VE_i_ is lower by R-fold (mostly because of lower IFR in re-infection and lower PI*).

### Values used and sensitivity analyses

For details on values used, justification (with supporting references), and sensitivity analyses’ ranges, see Appendix 1: Supplementary Methods. In brief, we use the 2021 world population pyramid age strata 0-19, 20-29, 30-39, 40-49, 50-59, 60-69, and 70 years and higher, dividing the last stratum further in community-dwelling (97%) and long-term care residents (3%). We assume that 10% in 0-19 years old, 20% in 20-29 years old, and 46% in higher age strata (overall 30% [sensitivity range 25-35%, retaining same age ratios]) had received at least one dose before any infection pre-Omicron. We assume that during Omicron, the remaining 56% of global population who remained uninfected by November 2021 were infected at least once until October 2024. An additional 18% of the global population were first vaccinated during Omicron with slightly less than a third (5%) receiving at least one dose before being infected.

We assume that absent vaccination all people would have been infected during the Omicron period. For pre-Omicron, we assume PI*=20% for all age strata by November 2021 (sensitivity range, 10%-40%) and that 5% of the population were first infected in the pre-Omicron period after vaccination.

For IFR in unvaccinated people pre-Omicron, we use estimates from a systematic review for non-elderly age strata; from meta-regressions for community-dwelling 70 years old and over; and from a meta-analysis of case-fatality rates and studies estimating asymptomatic infection rates for long-term care residents. Sensitivity range is informed by the same sources. Omicron IFR among unvaccinated is assumed to be one-third of pre-Omicron values.

We assume VE=75% (sensitivity range, 40-85%) pre-Omicron and 50% (sensitivity range, 30-70%) during Omicron. For people vaccinated after at least one infection, we assume R=5 (sensitivity range, 2.5-10).

For LE, the UN population division life table for 2021 (World, both sexes) is used taking the mid-point in each age bracket. For 70 years and above, LE at age 77 is considered for community-dwelling individuals and 2 years for long-term care facility residents. The main analysis considers f=0.5 for all strata (sensitivity range, 0.25-0.8).

### Numbers needed to treat

We calculated the number of vaccine doses required to avert one death and to save one life-year by dividing estimated benefits by the total number of 13.64 billion vaccine doses administered worldwide (https://data.who.int/dashboards/covid19/vaccines).

## RESULTS

### Lives saved

Table 1 shows the characteristics of the different strata used in the calculations (see Appendix 1: Supplementary Methods).^9–19^ In the main analysis (Table 2), 2.533 million lives were estimated to have been saved. They were mostly among people who were vaccinated before any infection (2.079/2.533, 82%). There were slightly more lives saved in the Omicron period (1.448/2.533, 57%). 89.6% of lives saved pertained to people 60 years and above. Children and adolescents accounted for only 0.01% of total lives saved and young adults 20-29 years old for another 0.07%.

**Table 1.** Characteristics of strata considered in the calculations.

| Age strata, years | World population | Proportion vaccinated before infection in pre-Omicron period (sensitivity range) | IFR in the pre-Omicron period (sensitivity range) |
| --- | --- | --- | --- |
| 0-19 | 2,664,996,463 | 0.1 (0.083-0.117) | 0.000003 (0-0.00002) |
| 20-29 | 1,209,691,398 | 0.2 (0.167-0.233) | 0.00002 (0-0.00007) |
| 30-39 | 1,173,183,969 | 0.46 (0.383-0.537) | 0.00011 (0.00005-0.00032) |
| 40-49 | 975,497,948 | 0.46 (0.383-0.537) | 0.00035 (0.00011-0.00077) |
| 50-59 | 849,924,808 | 0.46 (0.383-0.537) | 0.00123 (0.00047-0.00220) |
| 60-69 | 597,651,319 | 0.46 (0.383-0.567) | 0.00506 (0.00208-0.00860) |
| 70-, community | 468,997,399 | 0.46 (0.383-0.567) | 0.018 (0.013-0.023) |
| 70-, long-term care | 14,505,074 | 0.46 (0.383-0.567) | 0.12 (0.10-0.25) |
| All | 7,954,498,378 | 0.30 (0.25-0.35) |  |

IFR: infection fatality rate. Population pyramid data are from (9), long-term care residents are assumed to be 3% of the 70 years and older stratum (10,11), proportion vaccinated before infection in pre-Omicron period makes assumptions that depend on (12-14), IFR are based on (15-19) and details on the justification of the assumptions and sensitivity analysis ranges can be found in the Supplementary Methods.

**Table 2.** Lives saved by COVID-19 vaccination according to time period (pre-Omicron, Omicron) and whether vaccination was given to previously uninfected or previously infected people.

| Age strata | Lives saved:<br>pre-Omicron,<br>previously<br>uninfected | Lives saved:<br>pre-Omicron,<br>previously<br>infected | Lives saved:<br>Omicron,<br>previously<br>uninfected | Lives saved:<br>Omicron,<br>previously<br>infected | Total lives saved<br>(% of total saved) |
| --- | --- | --- | --- | --- | --- |
| 0-19 | 112 | 17 | 133 | 36 | 299 (0.01) |
| 20-29 | 677 | 104 | 806 | 220 | 1,808 (0.07) |
| 30-39 | 8,311 | 1,274 | 9,894 | 2,704 | 22,183 (0.9) |
| 40-49 | 21,988 | 3,371 | 26,176 | 7,155 | 58,690 (2.3) |
| 50-59 | 67,324 | 10,323 | 80,148 | 21,907 | 179,702 (7.1) |
| 60-69 | 194,753 | 29,862 | 231,849 | 63,372 | 519,836 (20.5) |
| 70-, community | 543,662 | 83,361 | 647,216 | 176,906 | 1,451,145 (57.3) |
| 70-, long-term care | 112,095 | 17,188 | 133,447 | 36,475 | 299,205 (11.8) |
| All | 948,922 | 145,501 | 1,129,669 | 308,776 | 2,532,869 (100) |

### Sensitivity analyses

Table 3 shows values ranging from 1.4 to 4.0 million lives saved in one-way sensitivity analyses. Benefits tended to be larger for the Omicron period, but not when R values were low or when pre-Omicron PI* was assumed to be large. The widest range for two-way sensitivity analyses was 1.0 to 6.0 million (considering lower and upper range for both VE and R).

**Table 3.** Results of sensitivity analyses for lives saved by COVID-19 vaccination.

| Previously infected/<br>Period | Lives saved<br>(millions) | Lives saved<br>(millions) | Lives saved<br>(millions) | Lives saved<br>(millions) | Lives saved<br>(millions) |
| --- | --- | --- | --- | --- | --- |
| | IFR<br>sensitivity<br>range | $D(\geq 1)_{\text{pre-omicron}}$<br>25-35% | PI* 10-40%<br>(pre-Omicron) | VE 40-85%<br>(pre-), 30-70%<br>(Omicron) | R 2.5-10 |
| No/Pre-omicron | 0.603-1.455 | 0.791-1.107 | 0.474-1.898 | 0.506-1.075 | 0.949 |
| Yes/Pre-omicron | 0.092-0.223 | 0.177-0.114 | 0.073-0.291 | 0.078-0.165 | 0.146 |
| No/Omicron | 0.717-1.732 | 0.941-1.318 | 1.130 | 0.678-1.581 | 0.565-2.259 |
| Yes/Omicron | 0.196-0.474 | 0.346-0.271 | 0.309 | 0.185-0.432 | 0.154-0.617 |
| Total | 1.608-3.884 | 2.256-2.810 | 1.986-3.628 | 1.447-3.254 | 1.814-3.971 |

### Life-years saved

In the main analysis, there were 14.8 million life-years saved, with sensitivity analyses ranging between 7.4 and 23.6 million life-years (Table 4). People over 60 years old accounted for most life-years saved (75.9%), but with very little contribution from long-term care residents (2% of total). 40-59 years old people also contributed a sizeable 20.6% of total. Children and adolescents (0.1%) and young adults 20-29 years old (0.3%) had negligible contributions.

**Table 4.** Estimates of life-years saved from COVID-19 vaccination.

| Age strata | Total Lives saved | Life expectancy, years | Life-years saved, f=0.5 (%) [main analysis] | Life-years saved, f=0.25 | Life-years saved, f=0.8 |
| --- | --- | --- | --- | --- | --- |
| 0-19 | 299 | 64 | 9,560 (0.1) | 4,780 | 15,297 |
| 20-29 | 1,808 | 49.9 | 45,114 (0.3) | 22,557 | 72,183 |
| 30-39 | 22,183 | 40.7 | 451,430 (3.0) | 225,715 | 722,288 |
| 40-49 | 58,690 | 31.8 | 933,167 (6.3) | 466,584 | 1,493,068 |
| 50-59 | 179,702 | 23.5 | 2,111,502 (14.2) | 1,055,751 | 3,378,403 |
| 60-69 | 519,836 | 16.2 | 4,210,672 (28.6) | 2,105,336 | 6,737,076 |
| 70-, community | 1,451,146 | 9.2 | 6,675,269 (45.3) | 3,337,635 | 10,680,431 |
| 70-, long-term care | 299,205 | 2 | 299,205 (2.0) | 149,603 | 478,728 |
| All | 2,532,869 |  | 14,735,921 (100) | 7,367,961 | 23,577,474 |

f: ratio of life expectancy in people dying from COVID-19 versus the total population in the same stratum.

### Numbers needed to treat

The overall benefit corresponds to one death averted per 5,400 vaccine doses (sensitivity range, one death averted per 3,500 to 9,300 vaccine doses) and 1 life-year saved per 900 (sensitivity range, 600 to 1,800) vaccine doses.

## DISCUSSION

We estimate that COVID-19 vaccination during 2020-2024 saved 2.5 million lives for 15 million life-years. For some perspective, 2.5 million lives correspond to approximately 1% of total global mortality in that period. Sensitivity analyses suggested ranges between 1.4 and 4 million averted deaths with 7.4-24 million life-years saved. However, uncertainty is substantially wider with multiple factors considered concurrently in multiple-way sensitivity analyses. The overall benefit corresponds to 1 death averted per 5,400 vaccine doses and 1 life-year saved per 900 vaccine doses. Numbers needed to treat may vary widely across age groups, given the steep age-risk gradient of COVID-19 fatality rates.

Lives saved during the Omicron period appeared slightly higher than those saved pre-Omicron. Estimated benefits during the Omicron period include both the benefits conferred by vaccination doses which were administered pre-Omicron (and maintained some protection during Omicron) and those conferred by vaccination first started or boosted in the Omicron period. Vaccinations started or boosted late in the pandemic may have contributed relatively little mortality benefit. Post-pandemic mortality benefits cannot be taken for granted; moving forward, optimizing COVID-19 vaccination recommendations would benefit from rigorous randomized trials.^20^ Moreover, the estimated Omicron period benefits become the minority under assumed large decreases in PI* x IFR x VE. The Omicron death burden was very low compared to earlier COVID-19 waves. This is unlikely to reflect mostly higher vaccination benefits. Pre-Omicron PI* carries large uncertainty since the extent of pre-Omicron viral spread depends on exposure load and measures taken. E.g., there was hardly any pre-Omicron viral circulation in China or New Zealand. Therefore, in these countries hardly any lives were directly saved by COVID-19 vaccination pre-Omicron; all benefit materialized in the Omicron period.

We estimate that 9 of 10 deaths averted and 8 of 10 life-years saved were in people 60 years old and over. While COVID-19 devastated long-term facilities,^21^ the proportion of life-years saved by vaccination in long-term facilities was only 2% of the total, mostly because of the very low LE of their residents. This may nevertheless vary across countries and institutions, depending on resident populations features (e.g. palliative care versus relatively healthy retired elderly).

The relative contribution of children, adolescents, and young adults to lives and life-years saved appears minimal. Assessment of absolute net benefits in these populations, if any, require careful consideration of potential additional benefits for non-lethal outcomes (e.g. hospitalizations and other symptomatic disease), as well as any deaths and other consequences from adverse effects (not included in our calculations).^22,23^ Cost-effectiveness ratios should be considered carefully in these age strata to document whether vaccination was worthwhile for younger vaccine recipients.^24^ The age groups 0-29 years old represent approximately half the global population. No worldwide data exist on how many vaccine doses were administered specifically in these age groups. However, if one-sixth of vaccine doses were given to these age groups, benefits would translate to 1 death averted per ∼100,000 vaccine doses. One might argue that vaccination of younger individuals may have diminished transmission to older, vulnerable individuals. However, vaccine effectiveness regarding infection risk was modest and very rapidly waning. False messaging that vaccination will substantially avert transmission may even have backfired. Risk compensation with increased exposure due to false reassurance may have resulted even in increased viral spread.^25^

Our estimates do not separate deaths averted from vaccine efficacy versus deaths caused from vaccination-related harms. Some may argue that, depending on risk aversion and regret considerations, a death caused because of harm may not carry the same weight as a death averted because of efficacy. Adverse events on COVID-19 vaccines remain a contentious topic. Randomized trial data are very limited.^26^ Estimates of risk from registries and other observations carry substantial uncertainty. However, as shown in Appendix 2, the number of deaths due to widely recognized and accepted adverse events (thrombosis, myocarditis, and deaths in highly debilitated nursing home residents) are probably approximately 2 orders of magnitude smaller than the overall benefit. Still, these harms may be important to weigh against benefits in specific sub-populations where there they have the highest frequency and where risk-benefit may change or even get reversed.

Our estimates include countries with very different pandemic and vaccination experiences. Of note, most non-high income countries (with the notable exception of China) had high proportions of their populations infected before vaccination.^13^ Given the prominent lack of global vaccine equity,^27,28^ probably only a minority of lives saved were in non-high income countries, even though they represent 84% of the global population. A modeling study estimated that 1.5 million lives could have been saved with universal vaccination against Omicron in low and low-middle income countries.^29^ Little of this benefit probably materialized. Inequity, inefficient vaccination campaigns, and vaccine hesitancy may have eroded substantially the benefit that might have been derived under ideal circumstances (see Appendix 3: “Deaths averted under ideal circumstances”).

Previous studies estimating lives saved by COVID-19 vaccination have focused on more limited time periods or more restricted areas, countries, or regions. The most-cited study to-date^3^ used modeling to estimate 14.4 million COVID-19 deaths and 19.8 million excess deaths averted across 185 countries in the first year of vaccination alone, with very limited uncertainty (13.7-15.9 million and 19.1-20.4 million for 95% credible intervals, respectively). These results vary markedly from our pre-Omicron period estimates. We did not estimate total excess deaths averted globally, as this is fraught with extreme uncertainties (30). However, for COVID-19 deaths, our results suggest over a log10-scale lower deaths averted by COVID-19 vaccination in that early period. Differences may be due to the unreliability of modeling in such complex circumstances^7^ and high estimates of IFR (especially in elderly) and VE (using short-term estimates available at that time) assumed in the modeling.^3^ Another modeling study estimated 620 thousand averted deaths from vaccination in the pre-Omicron period, increasing to 2.1 million based on underreporting assumptions.^4^ Our pre-Omicron estimates lie between these two estimates. Another study^5^ covered 34 countries/territories in Europe and estimated 1.6 million lives saved until March 2023 with 96% of lives saved among those 60 years or older and 60% during the Omicron period. The analyzed countries include approximately half of the global population of high-income countries. While we did not obtain estimates limited to these countries, our global estimates seem modestly more conservative. Differences may be due to implied IFR and VE estimates. However, we agree that the vast majority of lives saved were in the elderly with a slight preponderance of lives saved in the Omicron period. A study covering Latin America and the Caribbean until May 2022 estimated 1.18 million deaths averted (sensitivity range, 0.61-2.61 million) with 78% among those 60 years and over and 62% in the Omicron period.^31^

Several caveats should be discussed. First, the full range of uncertainty is larger than the range that we observe in presented sensitivity analyses. If one were to consider all factors in sensitivity analyses, the range of estimates would spread further. Moreover, our IFR estimates are derived from national seroprevalence studies before vaccination. For unvaccinated individuals, IFR in the second year (the pre-Omicron period that matters for calculation of lives saved) may have been lower with some effective treatments (e.g. dexamethasone) becoming available, better organization of healthcare services, and more experience in managing severe COVID-19. Moreover, there is debate on whether Delta was more or less lethal than the dominant variants of 2020.^32,33^

Second, for most factors considered, data informing their values come mostly from high-income countries. The picture is more uncertain in other countries. The two largest countries, China and India, have major uncertainty on estimates of COVID-19 disease burden,^34,35^ let alone vaccine benefits.

Third, VE assumptions try to amalgamate many different vaccines (of variable effectiveness,^36,37^ different doses, and different vaccination policies, along with waning effectiveness over time. Unavoidably these assumptions simplify very complex backgrounds. Analyses of VE based on observational data carry substantial uncertainty and bias.^38,39^ Healthy vaccinee bias is often observed,^40^ but it is difficult to adjust properly for its presence.

Fourth, life-year calculations are a contentious topic. A previous study that calculated adjusted LE in COVID-19 deaths based on comorbidities found small LE reduction versus the general population^41^ but was limited by incomplete information on comorbidities and their severity. Thus, there was probably substantial underestimation of the LE difference between COVID-19 victims and the general population. Another study showed that if LE reduction is modeled through a standardized mortality ratio (SMR) for COVID-19 victims versus the general population, mean LE at COVID-19 death in developed countries decreased from ∼10-12 years to ∼6-8 years with SMR=2,^41^ close to what our main analysis anticipates for f=0.5. That same study also estimated only 3.3-4.4 average discounted quality adjusted life-years.^42^ The SMR approach corresponds to higher f in young ages and larger f in elderly deaths; however, the share of life-years saved accounted by the elderly would be only slightly decreased.

In principle, if a disease/condition/event kills anyone regardless of health status, e.g. a nuclear bomb, then f=1; conversely, for a condition that appears exactly when a patient is dying from other co-existing ailments, f=0. The exact positioning of COVID-19 in that spectrum^43^ and the relative share of over- and under-counting of COVID-19 deaths^44^ are still debated with substantial consequences for estimated disease burden and vaccination benefits. Regardless, taking LE at age of death directly as a measure of anticipated life-years may lead to grossly misleading inferences.^45^ E.g. average LE at age of death for all death causes in western countries is ∼9-12 years anyhow^45,46^ – very close to the average unadjusted LE at age of death for COVID-19 deaths. Interestingly, if many people saved from COVID-19 death by vaccination had indeed limited LE, postponement of death would be short-lived. Such short-lived postponement may explain in part why substantial excess deaths were seen^47^ in several high-income countries in 2022-2023 despite high levels of vaccination. Of note, simple temporal correlations of excess deaths and vaccine use should not be used naively to infer vaccine effects. Vaccines may be used more extensively just before or during periods of higher viral circulation and death risk; this does not mean that they cause these deaths.

Finally, one may put COVID-19 vaccination benefits in perspective along with benefits from other available vaccinations. Comparisons should be cautious, given the different calculation methods used and acknowledging that mathematical models for other vaccinations may also not be fully reliable. However, one study^48^ estimated that vaccination for 10 pathogens across 112 countries in 2000-2019 saved 50 million lives; another 47 million may be saved in 2020-2030. Disability-adjusted life years saved were 2700 million and 2300 million, respectively. If these calculations are sound, COVID-19 vaccination apparently saved during 2020-2024 fewer lives than measles or hepatitis B vaccination in the same period, but more than vaccination for each of the other 8 pathogens. However, life-years saved by COVID-19 vaccination for the same period were more than 30-fold lower than the life-years saved from measles vaccination, 10-fold lower than from hepatitis B vaccination and substantially lower also than the life-years saved from HPV, yellow fever, *H. influenzae*, *S. pneumoniae*, and rubella vaccination.^48^ Therefore, even though COVID-19 vaccines are clearly a major achievement, their benefits do not necessarily match the benefits of several other widely used vaccines. Decrease in trust and increased hesitancy for these vaccines may be devastating.^49,50^ The COVID-19 pandemic and pandemic response created a more challenging landscape on how to overcome general vaccine hesitancy.^50–52^

In conclusion, COVID-19 vaccination offered major benefits during 2020-2024.

However, our estimates are substantially more conservative than early modeling efforts to calculate lives saved based on the first year of vaccination alone and strong assumptions on IFR and VE (3). Moreover, from our estimates the vaccination benefits seem to have been largely limited to the elderly portion of the global population. Long-term outcomes in both vaccinated and unvaccinated people should also be examined with further follow-up.

## Data Availability

All data are in the manuscript

## Acknowledgment

The work of John Ioannidis is supported by an unrestricted gift from Sue and Bob O’Donnell to Stanford University.

## Contributions

J.P.A.I. had the original idea, developed the new methods, performed analyses, and wrote the manuscript. A.M.P. and A.C. discussed concepts, collected background data and previous studies, interpreted analyses, and commented on the manuscript. S.B. discussed concepts, interpreted analyses, and commented on the manuscript. All authors have approved the final version.

## SUPPLEMENTARY MATERIAL

## APPENDIX 1

### SUPPLEMENTARY METHODS

#### General principles

The analysis compares the outcomes of global COVID-19 vaccination strategies to a counterfactual scenario of no vaccination. In calculating our main estimates, we do not consider separately deaths and other consequences from adverse effects of SARS-CoV-2 vaccines, nor do we make any adjustment for the quality of life-years saved. Moreover, we do not attempt to calculate indirect effects of COVID-19 vaccination which may have modulated excess deaths through an impact on non-COVID-19 causes of death.

#### Values used and sensitivity analyses

Population: The world population pyramid in 2021 was used in the calculations (1) considering age strata 0-19, 20-29, 30-39, 40-49, 50-59, 60-69, and 70 years and higher. The last age stratum was further divided into those living in the community and those living in long-term care facilities, assuming that the latter represent a 3% fraction of that age stratum; data on the size of this fraction are available mostly from high-income countries and there is sparse use of long-term care facilities in other countries (2,3). Residents of long-term care facilities are 0.7% of the total population in European countries, and 0.4% in the USA (2,3), while our assumption translates to ∼0.18% of the global population.

Proportion vaccinated before/after infection and before/after Omicron: By mid-November 2021, 53% of the global population had received at least one dose and 41% had been fully vaccinated with substantial disparity per income group (4). A systematic review has estimated (5) that 44% of the global population had been infected by mid-November 2021. If vaccination and prior infection were independent, then D(≥1)_pre-omicron_=53%x(1-0.44)=30% had received at least one dose before any infection (and similarly D(≥2)_pre-omicron_=0.41%x0.56=23% had been fully vaccinated before any infection) before the advent of Omicron. Vaccination and prior infection at a global level may be nearly independent events because the vast majority of infections were undetected, thus they would not have affected vaccination decisions. Moreover, most countries introduced campaigns that advised vaccination regardless of prior infection status.

We also assumed that D(≥1)_pre-omicron_ differed across age strata. Data are available mostly from some high income countries (6), showing substantially or far lower vaccination rates in children and young people in many countries. At global level, we assumed this age gradient may be more pronounced: 10% in those 0-19 years old, 20% in those 20-29 years old, and otherwise similar (46%) in higher age strata (overall 30%). In sensitivity analysis, we considered D(≥1)_pre-omicron_ ranging from 25% to 35%, retaining the same ratios between age strata as in the main analysis.

We assumed that after Omicron emerged, the remaining 56% of the global population who had not been infected by November 2021 were infected at least once until October 2024. People remaining uninfected are probably less than 5% in high-income countries with the most extensive vaccination programs (7) and probably overall very rare globally.

Calculations assuming 2% of the global population not infected until October 2024 did not materially change results (not shown). Between December 2021 and October 2024, an additional 71%-53%=18% of the global population received at least one dose and an additional 65%-41%=24% reached full vaccination status (6). Given the massive, rapid onslaught of the Omicron waves and the relatively slowing of vaccination of previously unvaccinated individuals after November 2021, we considered that only an additional D(≥1)_omicron_=5% (slightly less than a third of the 18%) were first vaccinated with at least one dose before being infected and assumed no distortion of the relative vaccination rates across age strata.

Proportion of people among those who are vaccinated who would have been infected in the absence of vaccination: PI* is a very uncertain parameter, because it largely depends on the strictness of other policies and measures taken at public and individual levels (e.g. lockdowns, masks, other restrictive measures or personal restrictions of exposure). We may assume that almost all people would have been infected during the Omicron period anyhow, since aggressive restrictive measures would have been impossible to hold for long. However, in separating the pre-Omicron period up to November 2021, several countries did achieve to have only minimal or even negligible circulation of the virus in that period. For the main analysis, we assumed for all age strata that PI*=20% of those vaccinated would have been infected until November 2021 in the absence of vaccination, during a mean follow-up of half a year of less. Sensitivity analyses considered values ranging from 10% to 40%. We also assumed that a small percentage of the population (5%) were infected for the first time in the pre-Omicron period after having received at least 1 vaccine dose, based on high vaccine effectiveness against infection in the first month but modest decrease by 6 months (8).

IFR: For IFR in unvaccinated people before the advent of Omicron, we used the median estimates obtained from a systematic review for non-elderly age strata and we used the interquartile range [IR] values in each age stratum for the sensitivity range (9). For the two youngest age strata the lower IQR value is 0% (due to no deaths observed in modest size population samples), but using instead the median or half the median for the lower sensitivity range yielded very similar total lives saved (not shown). For the 70 years old and over community-dwelling population we considered IFR=1.8% based on a previous meta-regression analysis that allows calculating expected IFR as a function of the proportion of people over 85 years old among those over 70 years (in the global population this proportion is 13.4%), and taking the median of estimates from two meta-regressions based on different seroprevalence study eligibility criteria (10). The estimates from the two meta-regressions were considered in the sensitivity analyses (range, 1.3 to 2.3). We considered IFR=12% for residents of long-term care facilities based on another review and meta-analysis (11) showing case-fatality rate of 22.71% from a summary of 51 studies covering 2020 and assuming approximately half of infections globally were missed because of being asymptomatic (given asymptomatic rates of 39-70% among infected residents in different surveillance studies) (12,13). In sensitivity analyses, we considered a range of 10% to 25%.

These estimates are derived from pre-vaccination data, but it is reasonable to assume a similar IFR for unvaccinated people during 2021. Conversely, for Omicron, we assumed that IFR was reduced to a third of the previous values in each stratum, as suggested by refs. (14,15).

Vaccine effectiveness for death: We assumed VE=75% during the pre-Omicron period and 50% during the Omicron period. This is an aggregate estimate considering the large heterogeneity of vaccination experiences (different vaccines, some of which had probably lesser effectiveness than others (16,17)), waning effectiveness especially with long-term follow-up (18), and also different vaccination experiences including many people who received only one or two doses in the pre-Omicron period. and one, two, three, or more doses in the Omicron period with overall lower effectiveness in the Omicron period). Sensitivity analyses considered VE values between 40% and 85% in the pre-Omicron period and 30% to 70% in the Omicron period.

R: For people vaccinated after at least one infection, we assumed R=5 (sensitivity range, 2.5 to 10), given the very low re-infection risk and lower IFR for them among previously infected in the pre-Omicron period (19), and the more common re-infections but with very low IFR in the Omicron period (20,21).

Life expectancy: For life expectancy, the UN population division life table for 2021 for World, both sexes (22) was used and for each age stratum, the mid-point in the age bracket was considered. For the 70 years and above, the life expectancy at age 77 was considered for the community-dwelling individuals and 2 years was used for long-term care facility residents based on epidemiological studies in such settings, e.g. (23,24).

For the factor f values, the main analysis considered f=0.5 for all strata, assuming that those who succumb to COVID-19, in the absence of infection would have half the life expectancy of the general population given the high burden of comorbidities and higher background socioeconomic burden in people dying from COVID-19 (25,26). In sensitivity analyses, values between 0.25 and 0.8 were considered.

Calculations were conducted in Excel. All key data used are in the manuscript and supplement and clarifications can be requested by the authors.

## APPENDIX 2

### DEATHS DUE TO ADVERSE EFFECTS OF VACCINES

Deaths due to adverse effects of vaccines have been a highly contentious and polarizing issue. Here, we present some estimates so as to put them into perspective against the potential estimated benefits due to vaccine efficacy. We focus on three types of adverse events for which there is strong consensus about their existence, even though there is still residual uncertainty about their exact frequency. Claims for potential long-term negative effects cannot be substantiated or refuted without longer-term follow-up with reliable data. Claims for large harmful effects on cardiovascular deaths early after vaccination, e.g. as suggested by a non-peer-reviewed analysis of the Florida Department of Health, have not been seen in other studies, some of which seem to have better study design. All evidence can be revisited, if more and better data and analyses accumulate.

The three very well documented harms that can result in death are thrombosis (along with thrombocytopenia) after adenovirus vector-based vaccines; myocarditis after mRNA vaccines (more common in young men); and death after vaccination in very debilitated nursing home residents.

For thrombosis, the risk is estimated as 20.3 per million doses in people 18-49 years old and 10.9 per million doses in older people (27) and mortality has ranged from 50% in early cases to 5% in later times with better recognition of the problem (28).

For myocarditis after mRNA vaccines, the risk is largely limited to men younger than 40 years old. According to (29), risk is seen in particular after a second dose and may be even higher with third doses of mRNA vaccines, and tends to be higher with Moderna than Pfizer vaccine. A living review suggests risks less than 150 per million in children and adolescents and much lower in higher ages (30). Death risk seems very low, with 1 death among 104 cases (1%) in a large series. A 4% mortality rate was seen in another series (31).

For deaths after vaccination in debilitated nursing home residents, data from Norway (32) suggest 10 likely and another 26 possible deaths among 29400 nursing home residents receiving the Pfizer mRNA vaccine, i.e. 0.4% to 1.2% mortality risk. It is unclear whether this applies also to other vaccines. See also ref. 33. Early VAERS data suggested a mortality rate of 53 per million people among all long-term care residents (34).

The total number of vaccine doses administered was 13.64 billion per the WHO dashboard. Detailed data on different vaccines exist for production until January 2022, when more than 11.5 billion doses had been produced (35). Allowing for the subsequent discontinuation of AstraZeneca and Johnson & Johnson adenovirus vector-based vaccines, we estimate that the administered vaccines included approximately 3.5 billion Pfizer mRNA, 1 billion Moderna mRNA, 3 billion adenovirus vector-based, and >6 billion inactivated and other vaccine doses.

Detailed data on age stratification world-wide for the administration of these vaccines are not available, but extrapolating from countries that have age-strata information (36), we use in the calculations here the following numbers: 1.5 billion adenovirus vector-based vaccine doses given in people <50 years old, 1.5 billion adenovirus vector-based vaccine doses given in older people, 0.5 billion mRNA doses given to men <40 years old, and 3 million doses given to very debilitated nursing home residents. Assuming death risks of 2 per million, 1 per million, 0.5 per million, and 0.5%, respectively, the estimated deaths are 3,000 for adenovirus vector-based vaccines in people <50 years old, 1,500 for adenovirus vector-based vaccines in older people, 500 for myocarditis, and 15,000 for deaths in very debilitated nursing home residents. The total amounts to 20,000 deaths. These estimates should be recognized as carrying very large uncertainty. Most importantly, recognition of these problems may have led to much lower utilization of these vaccines in high-risk populations, but the exact required adjustment is unknown.

In another approach one may consider data on all deaths attributed to COVID-19 vaccines in national data. Using national data from Qatar (37), among 6,928,359 doses administered, 138 deaths occurred within 30 days of vaccination; eight had a high probability (1.15/1,000,000 doses) and 15 had intermediate probability (2.38/1,000,000 doses) to be related to COVID-19 vaccination. For a total of 13.64 billion doses globally, the corresponding number of deaths would be 16,000 (high probability) to 48,000 (high or intermediate probability). These estimates, however, cannot adjust for differences in the Qatar versus global population demographics and vaccination experience.

Based on these data, deaths due to vaccination adverse events may have been about ∼100-fold lower than deaths averted for vaccination in the overall global population. However, the difference may have been much smaller or even reversed in some population subgroups at high risk of these adverse events.

## APPENDIX 3

### DEATHS AVERTED UNDER IDEAL CIRCUMSTANCES

Calculating formally the number of deaths that could have been averted by COVID-19 vaccination under ideal circumstances is highly speculative. The results may vary substantially under different assumptions about the feasible speed of vaccination campaigns and the eventual vaccination coverage achieved – perfect coverage is not a realistic expectation. However, the large majority of the benefit was derived when vaccinated before any SARS-CoV-2 infection and we assume in the main analysis that 35% of the global population overall and 51% of the elderly (that account for the vast majority of the benefit) were vaccinated before any infection (46% in the pre-Omicron phase and another 5% in the Omicron phase). Therefore, it is likely that close to half of the maximum achievable vaccination benefit in terms of lives saved materialized. The proportion is likely higher in high-income countries and lower in poor countries that suffered from the consequences of vaccine inequity.

As an illustrative calculation, one may assume the scenario to have the entire global population vaccinated already during the pre-Omicron period. One may assume that even under the most optimistic conditions, only 75% of the population would be vaccinated before any SARS-CoV-2 infection (as many had already been infected when vaccines became available and some more would be infected while deployment occurred, even if this were very fast). Then in the pre-Omicron period 75% are assumed to be vaccinated before infection and 25% are assumed to be vaccinated after infection; and in the Omicron period 70% have been vaccinated before infection and 30% have been vaccinated after infection. If vaccine choice and dosing were also optimized, we can then also assume the upper limits of the range of VE. Under these idealized circumstances, we estimate that 4,623,154 deaths would have been averted (1,881,231 in pre-Omicron/not infected, 125,415 in pre-Omicron/previously infected, 2,409,942 in Omicron/not infected, and 206,566 in Omicron/previously infected). The actual averted deaths (2,532,869) are therefore estimated to be 55% of the total that could have been averted under these idealized circumstances.

